# Performance of three rapid antigen tests against the SARS-CoV-2 Omicron variant

**DOI:** 10.1101/2022.02.17.22271142

**Authors:** Sanjat Kanjilal, Sujata Chalise, Adnan Shami Shah, Chi-An Cheng, Yasmeen Senussi, Rockib Uddin, Vamsi Thiriveedhi, Ha Eun Cho, Seamus Carroll, Jacob Lemieux, Sarah Turbett, David R Walt

## Abstract

Rapid antigen detection tests (RADTs) for severe acute respiratory syndrome coronavirus 2 (SARS-CoV-2) are now in widespread use in the United States. RADTs play an important role in maintaining an open society but require periodic reassessment to ensure test performance remains intact as the virus evolves. The nucleocapsid (N) protein is the target for the majority of RADTs and the SARS-CoV-2 Omicron variant has several N protein mutations that are previously uncharacterized. We sought to assess the impact of these mutations by testing 30 Omicron variant samples across a wide range of viral loads on three widely used RADTs: the iHealth COVID-19 Antigen Rapid Test, the ACON Laboratories FlowFlex COVID-19 Antigen Home Test, and the Abbott BinaxNOW COVID-19 Antigen Card, using 30 Delta variant samples as a comparator. We found no change in the analytic sensitivity of all three RADTs for detection of Omicron versus Delta, but noted differences in performance between assays. No RADT was able to detect samples with a cycle threshold (Ct) value of ≥27.5 for the envelope gene target on the Roche cobas RT-PCR assay. Epidemiologic studies are necessary to correlate these findings with their real-world performance.

## Introduction

Diagnostic testing for infection by severe acute coronavirus syndrome 2 (SARS-CoV-2) remains a cornerstone of efforts to control the coronavirus disease 2019 (COVID-19) pandemic. The reliance on centralized laboratory-based testing has eased with the introduction of rapid antigen tests, which are now available over-the-counter in the US or provided by the federal government. These assays can be self-administered, require little to no equipment and provide results within 15 minutes. The ability to test at the point of care with the onset of symptoms or prior to gatherings places them in a key role for maintaining an open society.

The SARS-CoV-2 nucleocapsid (N) is the most abundant protein expressed by the virus^1,2^ and is the target of the majority of rapid antigen detection tests (RADTs). Detection of the analyte is achieved through recombinant antibodies conjugated to gold nanoparticles that target specific epitopes on the N protein. The antigen-antibody complexes are carried by capillary action to a second set of antibodies which immobilize and concentrate the nanoparticles, making them visible to the naked eye. Mutations in the N protein have been previously described to cause decreases in antigen test sensitivity^3^, therefore periodic reassessment of test performance is necessary as new variants arise. The Omicron variant is characterized by a mutation (P13L) and a deletion (Δ31-33) near the N-terminal domain and two mutations adjacent to each other in the linker domain (R203K and G204R)^4^. We sought to characterize the impact of these newly described mutations on the analytic sensitivity of three widely used RADTs for at-home testing: the BinaxNOW COVID-19 Antigen Card (Abbott, Scarborough, Maine), the iHealth COVID-19 Antigen Rapid Test (Sunnyvale, California), and the FlowFlex COVID-19 Antigen Home Test (ACON Laboratories, San Diego, California), using their performance versus the Delta variant as a comparator.

## Methods

We collected 30 samples positive for the Omicron variant and 30 samples positive for the Delta variant from patients presenting to the Massachusetts General Hospital for clinical care between November 30 2021 and December 27 2021, except for nine Delta samples which were obtained between August and November 2021. Samples were collected from the anterior nares of patients, placed in universal transport medium and run on the cobas SARS-CoV-2 RT-PCR assay (Roche diagnostics, Pleasanton, California), which targets regions of the envelope (*E*) and *ORF1ab* genes. Variant calls were made using a combination of the TaqPath COVID-19 Combo Kit (ThermoFisher, Waltham, Massachusetts) to assess for spike gene target failure (SGTF), a proxy for the Δ69-70 spike mutation, and the TaqMan SARS-CoV-2 Mutation Panel (ThermoFisher, Waltham, Massachusetts), which amplifies a set of 6 spike protein mutations that characterize major variants of concern, including the Omicron and Delta variants. One sample was positive for only one target (K417N) due to having very low amounts of nucleic acid, but was assumed to be Omicron as it also had SGTF. All other samples were verified using multiple targets from the mutation panel. For each variant, we chose 10 samples with *E* gene cycle threshold (Ct) values of < 20, 10 samples with Ct values between 20 and 30 and 10 samples with Ct values > 30. Samples were not heat-inactivated.

For each RADT, we mixed the kit-supplied swab with 50 μL of sample for 15 seconds and then followed each assay’s instructions for use. Samples underwent one freeze-thaw cycle prior to examination on the iHealth assay and two freeze-thaw cycles for all other assays. Assays were run in duplicate for each RADT and results were evaluated after a 15 minute incubation period by two independent readers blinded to the variant status and Ct value of the sample. Samples were run for all three RADTs over a 2 day period. Chi-squared tests were used to compare the distribution of results by variant for a given RADT and logistic regression was used to estimate the impact of variant and RADT on the likelihood of test positivity after controlling for Ct values. This study was deemed non-human subjects research and approved by the Mass General Brigham Institutional Review Board (protocol 2021P003604).

## Results

For all three RADTs, there were no significant differences in the analytic sensitivity for the Omicron variant relative to the Delta variant. Table 1 shows the proportion of samples positive by Ct value range, RADT and variant.

**Table 1:** Proportion of tests positive by Ct value range, RADT and variant. There were no statistically significant differences in the proportion of tests positive by variant for any of the three RADTs overall, nor within the three Ct value ranges.

| Ct value range and RADT | n/total (%) positive |  |
| --- | --- | --- |
|  | Delta variant | Omicron variant |
| Overall |  |  |
| Abbott BinaxNOW | 15/30 (50%) | 13/30 (43%) |
| ACON FlowFlex | 15/30 (50%) | 15/30 (50%) |
| iHealth COVID-19 Antigen | 17/30 (57%) | 17/30 (57%) |
| ≤20 |  |  |
| Abbott BinaxNOW | 9/10 (90%) | 10/10 (100%) |
| ACON FlowFlex | 10/10 (100%) | 10/10 (100%) |
| iHealth COVID-19 Antigen | 10/10 (100%) | 10/10 (100%) |
| 20 - 30 |  |  |
| Abbott BinaxNOW | 6/10 (60%) | 3/10 (30%) |
| ACON FlowFlex | 5/10 (50%) | 5/10 (50%) |
| iHealth COVID-19 Antigen | 7/10 (70%) | 7/10 (70%) |
| ≥30 |  |  |
| Abbott BinaxNOW | 0/10 (0%) | 0/10 (0%) |
| ACON FlowFlex | 0/10 (0%) | 0/10 (0%) |
| iHealth COVID-19 Antigen | 0/10 (0%) | 0/10 (0%) |

Table 2 shows median Ct values and interquartile ranges by test result and the range of Ct value overlap between negative and positive tests, stratified by RADT.

**Table 2:** Median Ct values and overlap in Ct values between negative and positive results by RADT and variant.

| RADT and result | Median Ct (IQR) |  | Overlap range (Ct) |
| --- | --- | --- | --- |
|  | Negative | Positive |  |
| Abbott BinaxNOW |  |  |  |
| Delta variant | 31.1 (5.5) | 19.2 (3.5) | 19.6 - 27.1 |
| Omicron variant | 33.8 (6.7) | 19.1 (2.3) | 22.7 - 25.4 |
| ACON FlowFlex |  |  |  |
| Delta variant | 31.1 (5.5) | 19.2 (3.2) | 25.7 - 27.1 |
| Omicron variant | 34.0 (6.6) | 19.1 (4.9) | 24.7 - 24.9 |
| iHealth COVID-19 Antigen |  |  |  |
| Delta variant | 31.8 (3.8) | 19.6 (3.2) | N/A |
| Omicron variant | 34.6 (2.2) | 19.7 (5.5) | N/A |

Figure 1 depicts the distribution of test results by RADT, variant and Ct value. For the BinaxNOW there was overlap between negative and positive results in the Ct 19 to 27 range. When excluding one negative Delta variant sample that was an outlier, the BinaxNOW overlap range shrinks to 22 to 27. For the FlowFlex there was overlap in positive and negative samples in the Ct 24 to 27 range, while for the iHealth COVID-19 Antigen Test there was perfect discrimination between negative and positive results using a Ct value threshold of 27.5. No RADT was positive for samples with an *E* gene Ct value of >27.5 on the cobas SARS-CoV-2 assay.

**Figure 1:**
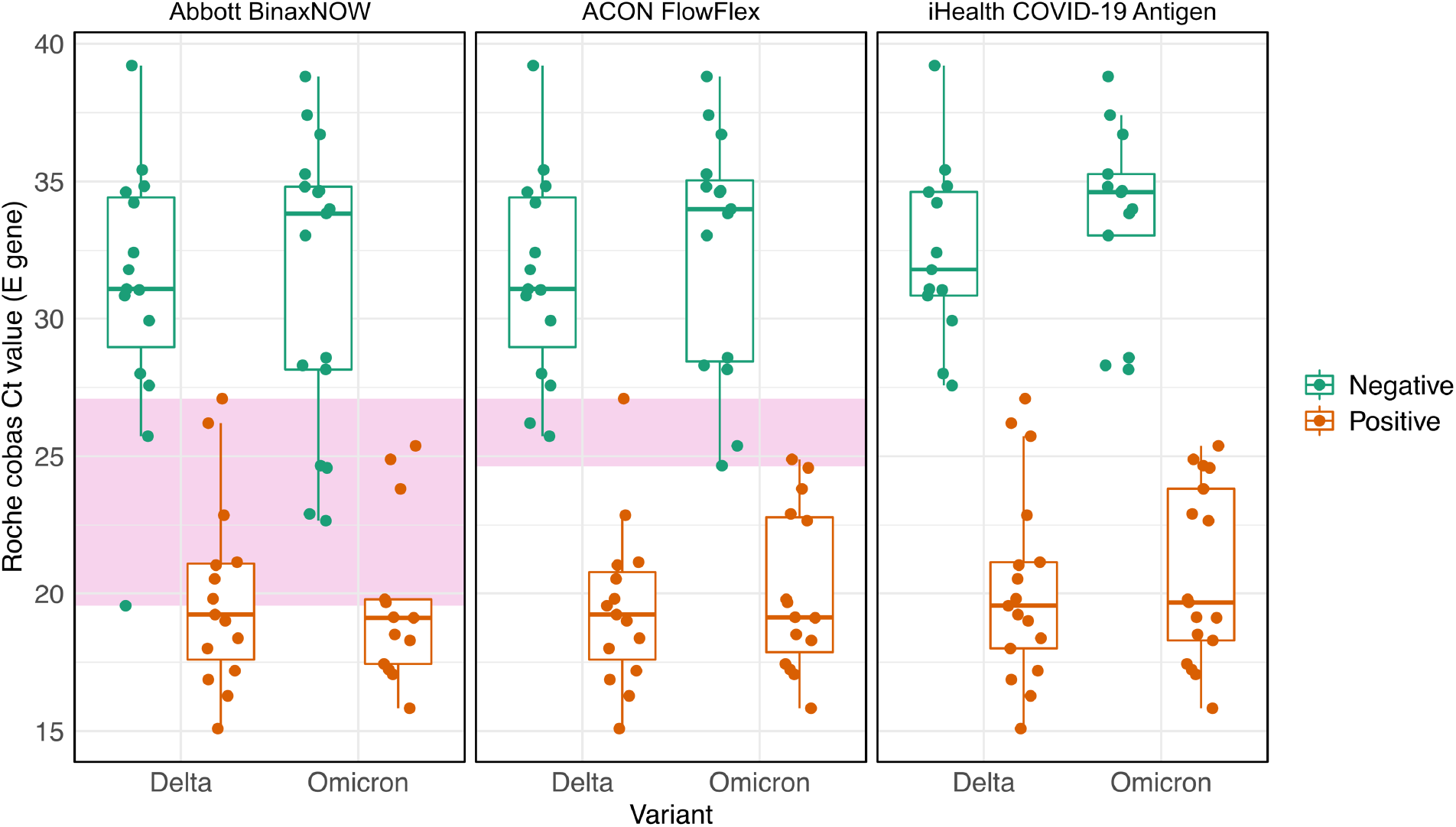
Distribution of Ct values by RADT, variant and test result. Boxes indicate median Ct and interquartile ranges. Shaded pink areas represent the range of Ct values for which there was overlap between negative and positive RADT test results. There was no overlap in positive and negative tests for the iHealth COVID-19 Ag test.

In multivariate analyses, variant type did not predict the odds of test positivity after controlling for Ct value and RADT, however the BinaxNOW had an 89% (95% CI 36% - 99%, p = 0.02) lower odds of being positive relative to the iHealth COVID-19 Antigen RADT. There was no statistically significant difference in the odds of being positive between the iHealth COVID-19 Antigen and the FlowFlex RADTs.

## Discussion

The emergence and spread of successive SARS-CoV-2 variants has come to define the COVID-19 pandemic over the past year and requires continual reassessment of vaccines, therapeutics and diagnostics. In this study, we show that three widely utilized RADTs for at-home diagnosis of SARS-CoV-2 infection continue to perform as expected despite a number of mutations in their target, the nucleocapsid protein. While there was no difference in the analytic sensitivity of RADTs between Delta and Omicron variant samples, we note differences in the performance by assay type. These results should be interpreted with caution given our relatively small sample size. We also note a non-significant trend towards decreased detection of the Omicron variant for the Abbott BinaxNOW, which we have noted in a prior study from our group using a different sample set^5^.

Three studies examining the analytic sensitivity of the BinaxNOW against the Omicron variant have shown no statistically significant changes in test performance^6-8^. However, most of these studies were either very small or did not compare Omicron and Delta samples side by side. A comparison of the Abbott Panbio COVID-19 Ag Rapid Test to other RADTs available on the Australian market using viral culture from a single Delta variant and Omicron variant sample showed equivalent sensitivities across a range of dilutions^6^. In contrast, a similar study by Bekliz et al using viral culture and paired clinical samples did find attenuated analytic sensitivity for detection of Omicron by the Panbio test, relative to the Delta variant^9^. The Panbio uses the same N protein epitopes as the BinaxNOW, which is marketed in the United States. The reasons for these differing results are not fully known, but they highlight the need for a repeat study using a larger set of samples with Ct values in the 20 to 30 range to resolve whether the BinaxNOW and Panbio assays have lower analytic sensitivity for Omicron.

The largest clinical study examining the BinaxNOW was performed at a community testing site in San Francisco during a time when rates of test positivity exceeded 40%^8^. The BinaxNOW was able to reliably detect positive samples up to a Ct of 30 for the PCR assay used in this study. While this value is higher than our detection threshold of 27.5 cycles for the *E* gene of the cobas SARS-CoV-2 RT-PCR assay, this could be explained by differences in reaction efficiencies, thresholding algorithms and a host of other factors as opposed to true differences in viral loads^10^. Direct comparison of Ct values between studies is challenging as distributions can have systematic biases across platforms.

No samples were RADT positive above a Ct value of 27.5 on our RT-PCR assay, but how this threshold relates to infectivity is an open question. The risk of SARS-CoV-2 transmission increases in direct proportion with the viral load of the index patient^11^ but is also dependent on the duration of exposure, the presence of masks, ventilation, host immunity, symptomatology and characteristics of the virus variant^12^. Infectivity is a multidimensional phenomenon that involves numerous variables operating on continuous scales, therefore it is difficult to assign a single Ct value as a threshold without additional data correlating results with contact tracing.

The iHealth COVID-19 test has been distributed to millions of US citizens through a free distribution program offered by the federal government. To our knowledge, this study is the first to independently evaluate the performance of this assay against the Omicron variant. While both the iHealth and FlowFlex RADTs performed well with our sample set, the iHealth test had a trend towards higher sensitivity and also had the best discrimination between positive and negative tests. The BinaxNOW had positive and negative results across an overlap of eight PCR cycles, corresponding to a 256-fold difference in the amount of virus present in a sample, assuming a reaction efficiency of 100%. The range of overlap for the FlowFlex was limited to 3 PCR cycles, which is an 8-fold difference in the amount of virus present in a sample. While the reasons for the overlap at these viral loads is not known, a significant number of people tested during an outbreak may have levels that fall within this range. Further study is necessary to understand whether this variation is reproducible in other contexts.

The major limitation of our study is the sample size, which limits drawing statistically significant conclusions regarding small differences in test performance. A larger study is warranted to further investigate the differences seen between our RADTs, as even small differences can have a large impact when scaled to the population level. Another limitation is our use of frozen samples in universal transport media rather than direct testing from a patient, but we would not expect there to be a major impact from one to two freeze-thaws on assay performance and the volume of analyte used in each assay was optimized in an earlier study for mimicking real-world performance^12^. A major strength of this study was the ability to compare three different RADTs using identical clinical samples, which allows for a robust comparison of performance.

In summary, the analytic sensitivity versus Omicron remains stable in our head-to-head comparison of three of the most common RADTs in use in the United States. However, there were differences in inter-assay performance that warrant further study. Our findings will provide a degree of assurance that at-home testing should perform as expected compared to prior waves and also sets a baseline for comparison with future SARS-CoV-2 variants.

## Data Availability

All data produced in the present study are available upon reasonable request to the authors

## Acknowledgements

This study was funded by a grant from the Massachusetts Consortium for Pathogen Readiness.

